# Artificial Intelligence for Early Detection and Prognosis Prediction of Diabetic Retinopathy

**DOI:** 10.1101/2025.03.29.25324873

**Authors:** Yudi Kurniawan Budi Susilo, Dewi Yuliana, Mahani Mahadi, Shamima Abdul Rahman, Azrin Esmady Ariffin

## Abstract

This review explores the transformative role of artificial intelligence (AI) in the early detection and prognosis prediction of diabetic retinopathy (DR), a leading cause of vision loss in diabetic patients. AI, particularly deep learning and convolutional neural networks (CNNs), has demonstrated remarkable accuracy in analyzing retinal images, identifying early-stage DR with high sensitivity and specificity. These advancements address critical challenges such as intergrader variability in manual screening and the limited availability of specialists, especially in underserved regions. The integration of AI with telemedicine has further enhanced accessibility, enabling remote screening through portable devices and smartphone-based imaging. Economically, AI-based systems reduce healthcare costs by optimizing resource allocation and minimizing unnecessary referrals. Key findings highlight the dominance of Medicine (819 documents) and Computer Science (613 documents) in research output, reflecting the interdisciplinary nature of this field. Geographically, China, the United States, and India lead in contributions, underscoring global efforts to combat DR. Despite these successes, challenges such as algorithmic bias, data privacy, and the need for explainable AI (XAI) remain. Future research should focus on multi-center validation, diverse AI methodologies, and clinician-friendly tools to ensure equitable adoption. By addressing these gaps, AI can revolutionize DR management, reducing the global burden of diabetes-related blindness through early intervention and scalable solutions.

## 1 Introduction

This comprehensive review explores the multifaceted role of artificial intelligence (AI) in the early detection and prognosis prediction of diabetic retinopathy (DR), emphasizing the importance of integrating advanced computational methods into clinical workflows to improve diagnostic accuracy and patient outcomes [1], [2], [3]. AI algorithms have been increasingly applied to analyze retinal images, leveraging both convolutional neural networks (CNNs) and deep learning techniques to identify subtle vascular changes and microaneurysms that are indicative of DR at its earliest stages [2], [4], [5]. The potential of these methods to assist ophthalmologists in reducing variability in image interpretation and improving screening throughput is underscored by the rapid advancements observed over the last decade, which have significantly decreased the rate of misdiagnosis while simultaneously enhancing prognostic capabilities [1], [3], [6]. In addition, the integration of AI with telemedicine solutions has enabled remote screening and monitoring in underserved areas, thereby addressing the growing disparity in healthcare accessibility [2], [7], [8]. These developments, coupled with cost-effectiveness and scalability, pose a disruptive shift in conventional DR management paradigms, promising to alleviate the socio-economic burden associated with diabetes and its ocular complications [9], [10], [11].

The global increase in diabetes prevalence has precipitated an urgent need for innovative diagnostic tools to mitigate the risk of vision loss due to DR, and AI-based systems have emerged as a viable solution to this mounting challenge [1], [10], [12]. Epidemiological studies indicate that early detection and prompt intervention can prevent visual impairment in over 90% of DR cases, a fact that underscores the clinical and public health significance of deploying AI in screening programs [13], [3], [6]. The rapid dissemination of diabetes worldwide, coupled with the high cost and limited availability of trained retinal specialists, necessitates the adoption of automated diagnostic systems that can efficiently analyze large volumes of retinal images [2], [7], [9]. Furthermore, AI systems have demonstrated remarkable sensitivity and specificity in detecting varying stages of DR, thereby providing clinicians with valuable secondary opinions and potentially reducing the human error associated with manual grading [14], [15], [16]. Consequently, the transformative potential of AI is not only evident in its diagnostic accuracy but also in its capacity to optimize resource allocation and enhance healthcare delivery at a systems level [9], [17], [18].

Diabetic retinopathy is a microvascular complication of diabetes that can lead to irreversible blindness if not diagnosed and treated in a timely manner, and the early identification of this condition is critical to preserving vision [1], [13], [4]. Traditional screening methods often involve labor-intensive manual analysis of fundus photographs and OCT images, a process that is both time-consuming and subject to interobserver variability [15], [19], [5]. AI-based systems, leveraging state-of-the-art image processing techniques, have been developed to overcome these limitations by automating the detection process and standardizing interpretation outcomes across diverse patient populations [2], [3], [20]. Recent studies have validated the clinical efficacy of these systems by demonstrating high concordance with human expert assessments in large-scale multicenter trials, thereby reinforcing their utility as an adjunct in routine clinical practice [21], [22], [23]. The incorporation of AI into DR screening protocols is poised to revolutionize patient care by ensuring earlier diagnosis, enabling timely therapeutic interventions, and ultimately reducing the incidence of diabetes-related blindness [17], [10], [8].

## 2 Literature

Recent advancements in deep learning and neural network architectures have significantly improved the capability of AI algorithms to detect early signs of DR from fundus imaging with unprecedented accuracy [2], [3], [20]. These benefits are particularly evident in convolutional neural network (CNN) approaches, where large annotated datasets enable the system to learn intricate retinal features that may elude even experienced clinicians [2], [4], [24]. The robust performance of these models is confirmed by high sensitivity and specificity metrics that have been consistently reported across clinical validation studies, suggesting that AI can reliably identify the subtle changes in retinal morphology that precede overt DR [14], [16], [3]. Moreover, the scalability of AI models allows for rapid screening of large patient cohorts while preserving high diagnostic performance, a factor that is critically needed in regions with limited access to ophthalmologic care [7], [8], [18]. As a result, the integration of AI for early detection not only enhances screening efficiency but also offers a promising pathway for disease monitoring and risk stratification over time [20], [9], [6].

A notable challenge in DR screening has been the intergrader variability that arises from the subjective nature of manual image interpretation, yet AI-based grading systems have demonstrated the ability to standardize diagnostics across different populations [15], [19], [3]. Variability in human grading has long been recognized as a barrier to consistent and timely diagnosis, leading to adverse patient outcomes and increased healthcare costs [15], [10], [25]. In contrast, validated automated deep learning software offers reproducible performance by applying uniform criteria to all images, thereby reducing diagnostic inconsistencies [15], [3], [26]. Clinical studies have highlighted that automated systems can achieve comparable or superior performance to human graders, especially in the identification of mild and early-stage DR, where variability is most pronounced [14], [15], [22]. This standardization is pivotal for large-scale screening initiatives that aim to reduce the burden on ophthalmic specialists while maintaining high diagnostic standards [3], [17], [18].

Telemedicine has emerged as an essential component of contemporary DR screening programs, particularly in rural and underserved regions where access to specialized care is limited [2], [7], [27]. The convergence of AI with telemedical systems allows for the remote acquisition and automated analysis of retinal images, which enhances both the speed and accuracy of DR detection [7], [16], [8]. By leveraging smartphone-based fundus cameras and portable imaging devices, AI systems can facilitate point-of-care screening even in resource-constrained settings, promoting earlier intervention for diabetic patients [16], [28], [27]. The decentralized nature of these technologies enables extensive population screening campaigns that bypass traditional healthcare infrastructure limitations [7], [10], [29]. Consequently, the synergy between telemedicine and AI represents a paradigm shift in the delivery of eye care, reducing geographic and economic barriers to effective DR management [2], [16], [8].

Economic evaluations of AI-based DR screening have consistently demonstrated that automated systems can significantly lower the overall cost of care compared to conventional human-based grading [14], [9], [11]. Studies indicate that AI solutions yield cost savings by reducing the workload on specialized personnel, optimizing resource allocation, and minimizing the frequency of unnecessary referrals [9], [17], [29]. The integration of automated screening tools has been shown to be particularly cost-effective in high-volume clinical settings, where the reduction in diagnostic turnaround times translates into better patient outcomes and lower system-wide expenditures [14], [9], [11]. In addition, by detecting DR at an earlier stage, AI-based screening can prevent the progression to advanced disease states that require more aggressive and costly interventions [1], [13], [6]. The economic advantages of AI also extend to its capacity to provide scalable solutions in regions where traditional screening methods are limited by a shortage of trained healthcare professionals [17], [30], [18].

The real-world implementation of AI in DR screening is exemplified by systems that have received regulatory approvals, such as the IDx-DR and EyeArt platforms, which are now integrated into clinical practice in several countries [4], [22], [6]. These systems have undergone rigorous clinical testing to ensure that their performance metrics meet or exceed those of manual grading by expert clinicians [4], [3], [6]. Their deployment in routine screening workflows has not only validated their diagnostic precision but also underscored the importance of comprehensive clinical validation studies involving diverse patient cohorts [22], [23], [17]. The ability of these platforms to process high-resolution retinal images rapidly and with high accuracy has made them instrumental in reducing the incidence of undiagnosed DR among diabetic populations [1], [3], [6]. Such advancements provide strong evidence in support of the widespread adoption of automated DR screening tools in both high- and low-resource healthcare settings [22], [17], [8].

## 3 Method

This study adopted a rigorous systematic bibliometric analysis following the PRISMA framework to investigate the application of artificial intelligence in diabetic retinopathy detection and prognosis prediction. The research methodology was designed to provide both quantitative and qualitative insights through multiple phases of data collection, screening, and analysis. We utilized Scopus as our primary database, focusing on journal articles published between 2022 and 2025 in computer science and medicine, to ensure we captured the most recent and relevant advancements in this rapidly evolving field. Our comprehensive search strategy incorporated three key thematic areas: AI methodologies, diabetic retinopathy terminology, and clinical applications, which yielded an initial pool of 5,878 records.

**Figure 1:**
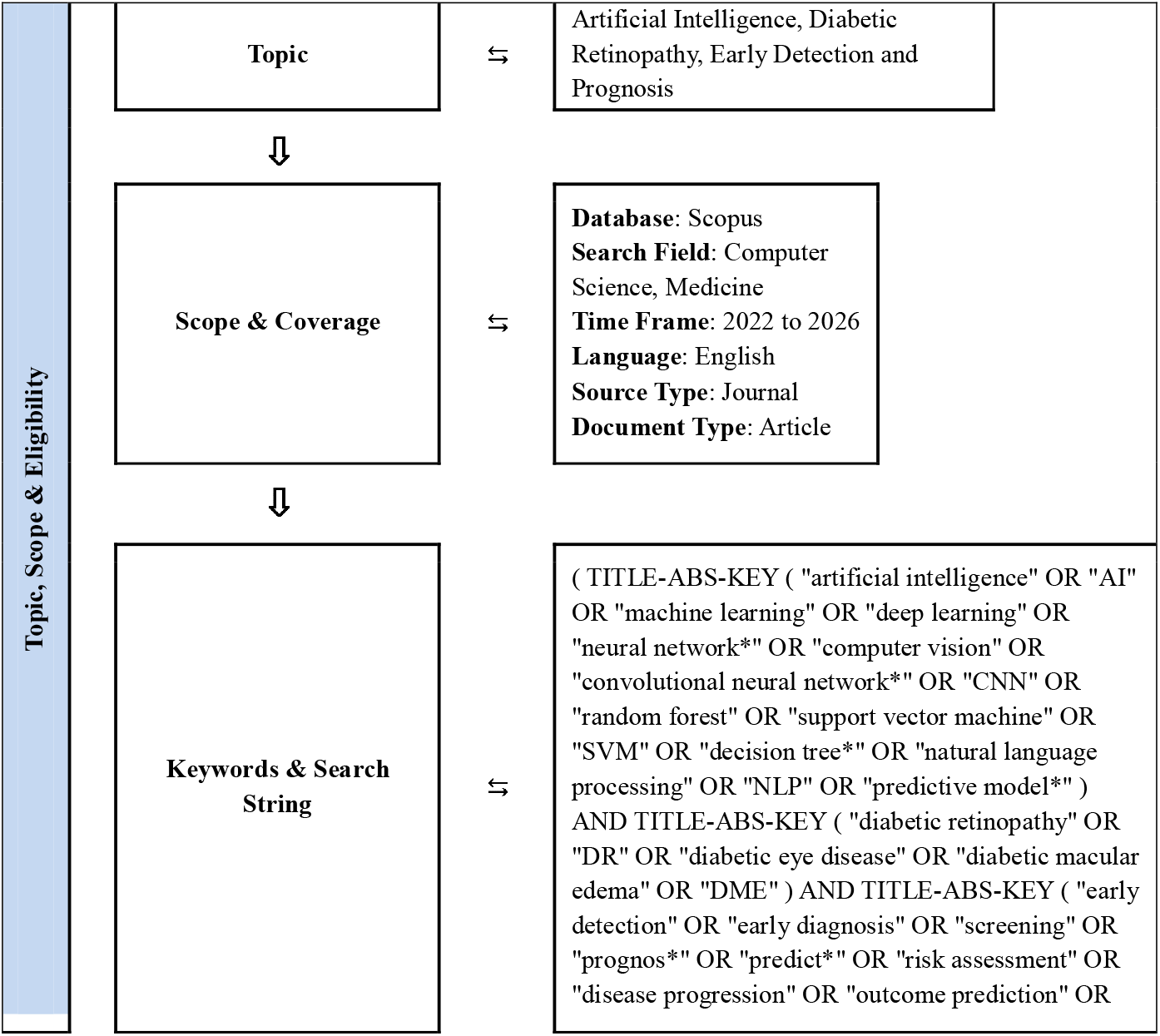
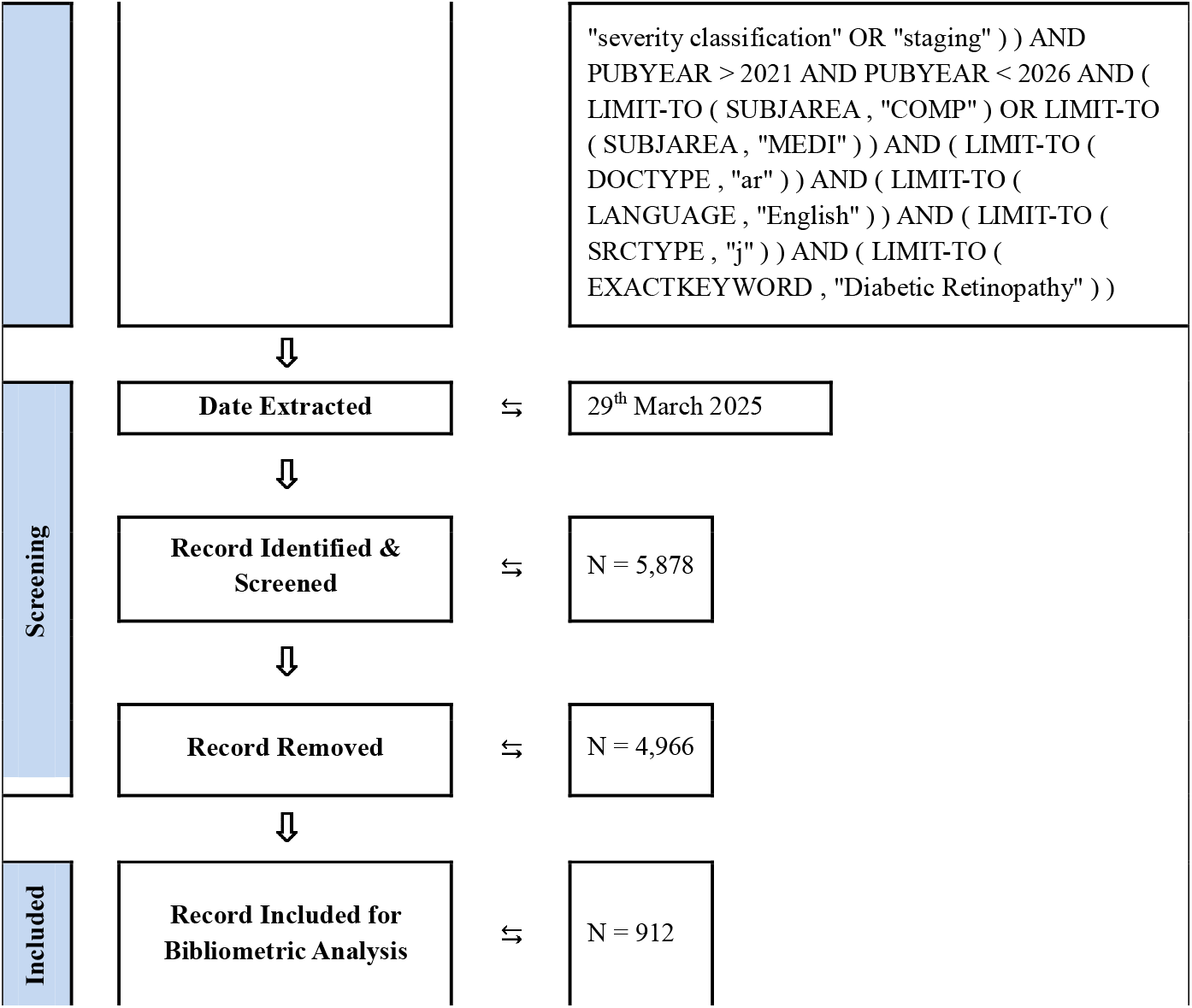
Flow diagram of search strategy for topic Artificial Intelligence, Diabetic Retinopathy, Early Detection and Prognosis from 2022 to 2025

The screening process followed strict PRISMA guidelines, beginning with duplicate removal before progressing through title/abstract screening and full-text evaluation conducted by independent reviewers. This meticulous approach resulted in 912 high-quality articles selected for in-depth analysis, with careful documentation of exclusion criteria at each stage to maintain transparency. For the bibliometric analysis, we employed a suite of specialized tools including VOSviewer for network visualization and science mapping, Harzing’s Publish or Perish for citation analysis, and Microsoft Excel for data organization and trend analysis. These tools enabled us to conduct performance analyses of publication patterns, science mapping of conceptual domains, and content analysis of AI techniques and clinical applications.

To ensure methodological rigor, we implemented several quality assurance measures such as inter-rater reliability checks, search string validation, and peer debriefing. Our analytical approach combined quantitative metrics like citation counts and author productivity with qualitative assessments of research themes and methodological quality. The visual representation of our methodology through the PRISMA flow diagram effectively communicated our systematic literature selection process, while the multi-dimensional analysis framework provided both macro-level patterns and micro-level insights. This comprehensive approach not only mapped the current state of knowledge in AI applications for diabetic retinopathy but also identified significant gaps and opportunities for future research, offering valuable insights for researchers and clinicians working at the intersection of artificial intelligence and ophthalmology.

## 4 Result

The analysis of research output by country reveals a distinct global distribution of scientific contributions. China leads with a substantial 487 documents, demonstrating its dominant position in this research domain, followed by the United States with 347 publications, reflecting the strong research ecosystems in these two nations. India ranks third with 279 documents, highlighting its growing expertise in both medical research and artificial intelligence applications.

**Figure 2.**
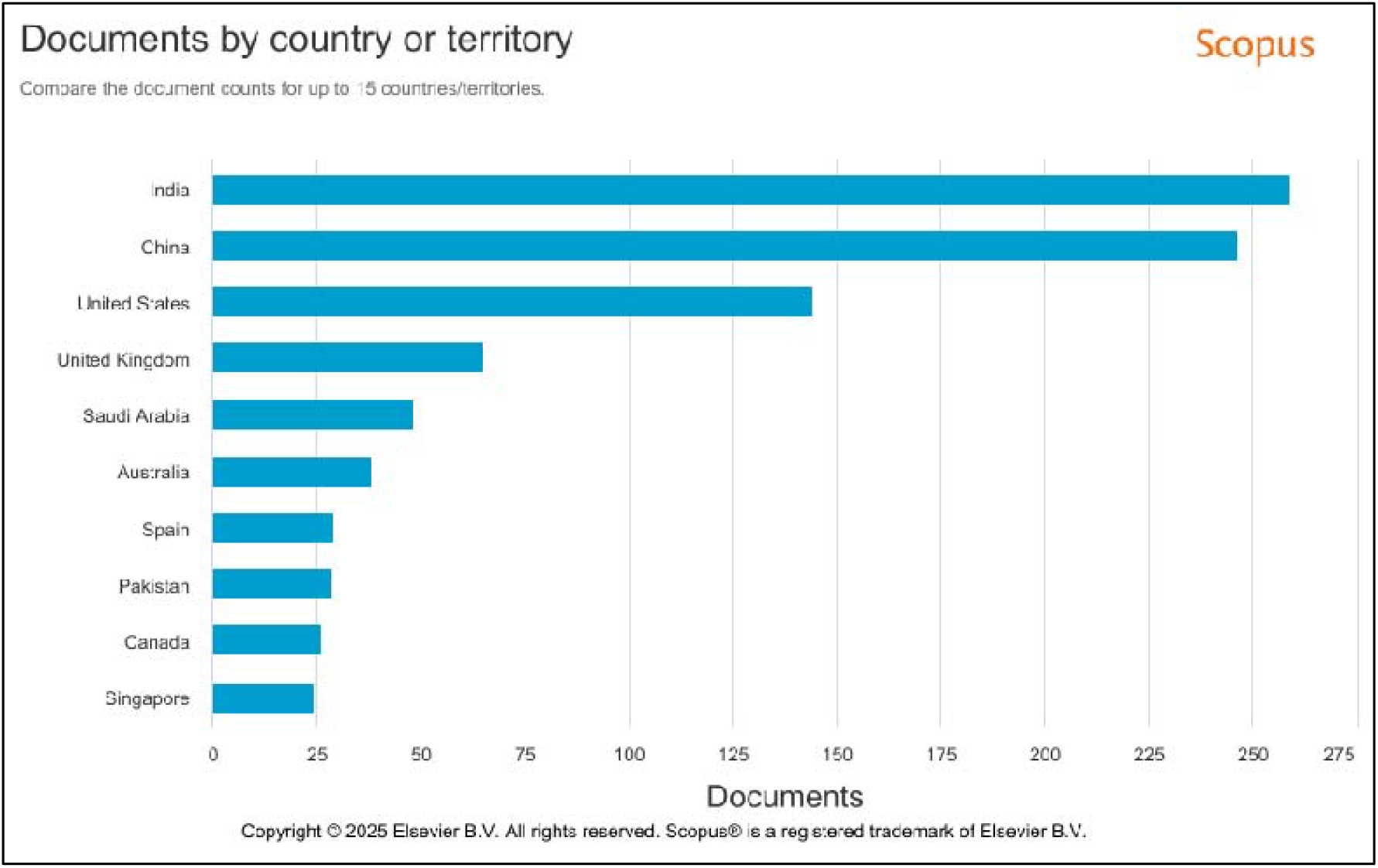
Documents published by country or territory between 2022 and 2025

The United Kingdom occupies fourth place with 137 publications, while Australia completes the top five with 91 documents. Notably, Asian countries collectively show strong representation, with Singapore and South Korea tied at 83 publications each, Japan contributing 67, and Hong Kong adding 49. The Middle East is represented by Saudi Arabia with 63 documents, while European contributions come from Germany (61), Spain (58), Italy (48), and Switzerland (44).

Pakistan’s inclusion with 40 documents indicates emerging research activity in this field. This geographical distribution suggests that research in AI for diabetic retinopathy is concentrated in countries with robust healthcare systems, strong technological infrastructure, and significant investments in medical AI research, while also showing promising growth in developing nations addressing high diabetes prevalence through technological solutions. The data underscores the global nature of this interdisciplinary research field, combining ophthalmology, diabetes care, and artificial intelligence innovation.

### 4.1 Documents by subject area

The subject area distribution of research reveals a clear interdisciplinary pattern, with Medicine (819 documents) emerging as the dominant field, accounting for nearly one-third of all publications. This strong medical predominance reflects the clinical orientation of diabetic retinopathy research, where AI solutions are primarily developed to address real-world diagnostic and prognostic challenges in ophthalmology. Computer Science (613 documents) follows as the second-largest contributor, highlighting the crucial role of machine learning algorithms, image processing techniques, and computational models that underpin these medical applications. The substantial representation of Engineering (534 documents) further emphasizes the importance of technological implementation, particularly in developing medical imaging systems and diagnostic devices.

**Figure 3.**
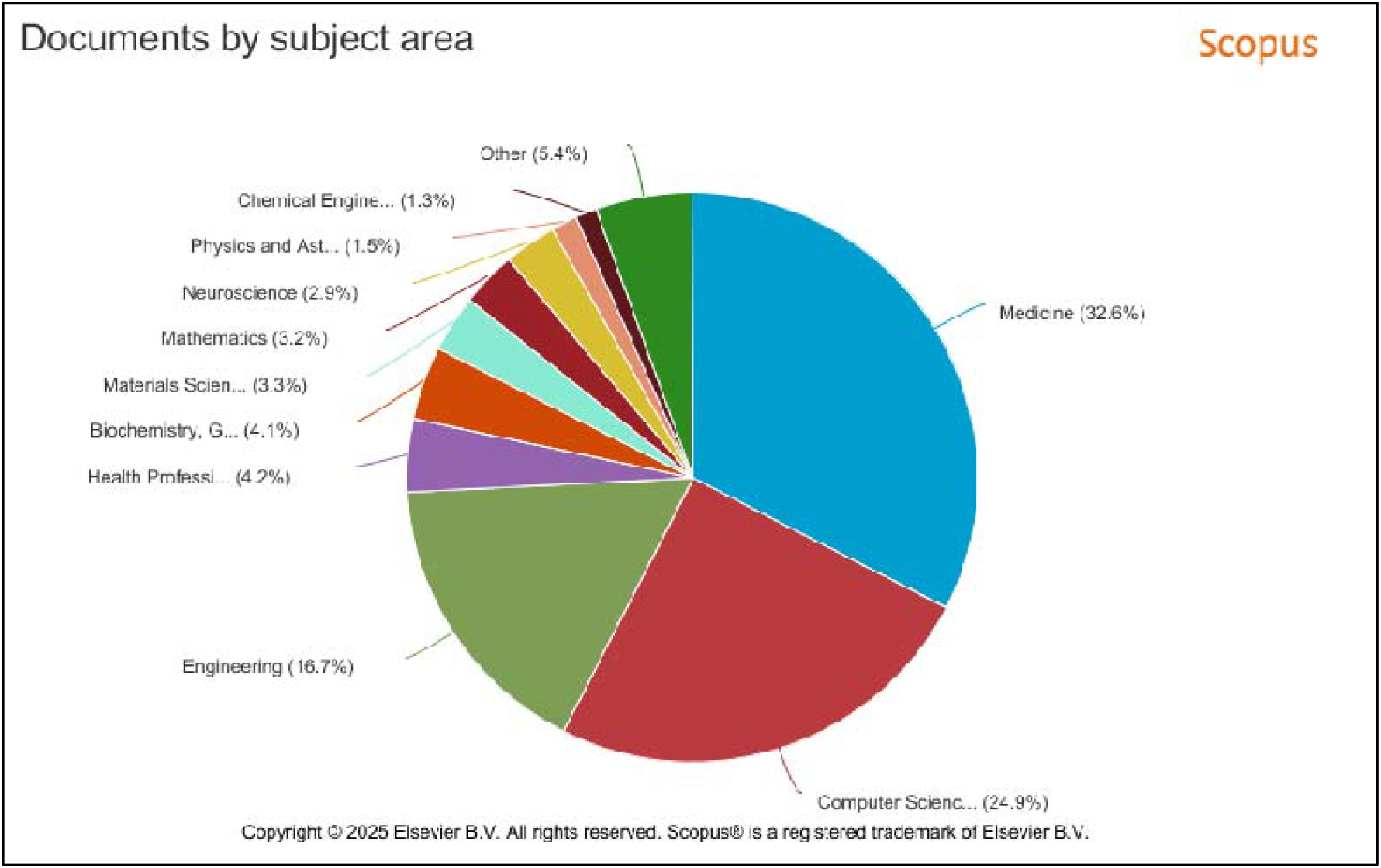
Documents published by subject area between 2022 and 2025

A secondary tier of disciplines demonstrates the breadth of this research field. Biochemistry, Genetics and Molecular Biology (187 documents) and Neuroscience (159 documents) likely represent studies exploring the biological mechanisms of diabetic retinopathy or neural network-inspired AI architectures. The presence of Physics and Astronomy (143 documents) may relate to advanced imaging technologies like optical coherence tomography, while Health Professions (126 documents) reflects translational research focused on clinical workflow integration. Materials Science (103 documents) and Chemical Engineering (57 documents) contributions probably concern sensor development or contrast agents for retinal imaging.

The long tail of other disciplines including Multidisciplinary (101), Mathematics (96), and Decision Sciences (20) illustrates how this field attracts diverse methodological approaches, from mathematical modelling of disease progression to decision support system development. Notably, the minimal representation of pure Chemistry (30 documents) suggests most research focuses on applied rather than fundamental science aspects. This distribution pattern confirms that AI applications for diabetic retinopathy represent a true convergence field, where medical needs drive technological innovation, and where successful solutions require close collaboration between clinicians, computer scientists, and engineers. The predominance of Medicine and Computer Science publications suggests that while the field welcomes diverse contributions, its core remains firmly anchored in clinical problem-solving supported by advanced computational methods.

### 4.1 Co-occurrence link between keywords

The keyword co-occurrence analysis reveals several important thematic clusters and research trends. The strongest connections emerge around core clinical and technical concepts, with “diabetic retinopathy” (total link strength=19) serving as the central node, reflecting its position as the primary medical focus of these studies. “Deep learning” (link strength=12) appears as the dominant AI methodology, significantly outpacing other technical approaches in this research space.

**Figure 3.**
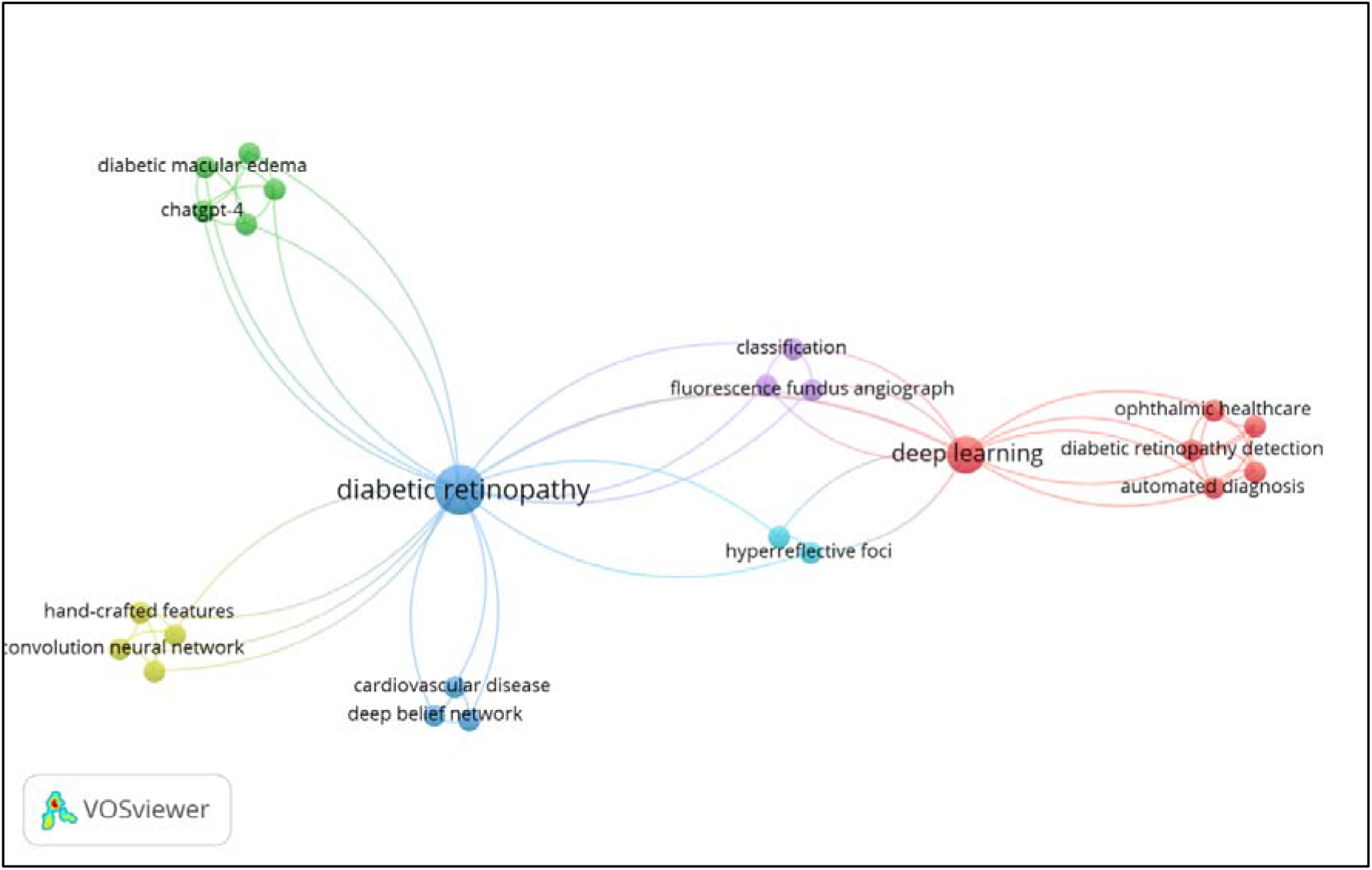
Network visualization of co-occurrence link between keyword for research topic Artificial Intelligence, Diabetic Retinopathy, Early Detection and Prognosis, between year 2022 and 2025.

Several noteworthy thematic patterns emerge from the mid-range connections (link strength=5). The cluster around “automated diagnosis,” “diabetic retinopathy detection,” and “medical image analysis” demonstrates the field’s emphasis on developing autonomous screening tools, while terms like “capsule network” and “convolution neural network” (strength=4) specify the preferred neural network architectures. The appearance of emerging concepts like “code-free prompt” and “no-code” suggests growing interest in making AI solutions more accessible to clinicians without programming expertise. Clinical applications are represented through terms such as “diabetic macular edema,” “ophthalmic healthcare,” and “risk calculator,” indicating a balance between technical development and practical implementation.

The presence of “classification” (strength=4) and “feature map” (strength=4) reveals ongoing methodological focus on image categorization and feature extraction techniques. The relatively equal strength of connections across technical (“capsule network”), clinical (“diabetic macular edema”), and implementation (“risk calculator”) terms suggests this research field maintains a healthy equilibrium between algorithm development and clinical translation. However, the dominance of “deep learning” over other AI approaches indicates a potential research gap regarding alternative machine learning methods in diabetic retinopathy applications. These patterns collectively portray an evolving research landscape where advanced neural networks are being actively adapted to address specific clinical challenges in diabetic eye disease, with growing attention to deployment-friendly solutions that bridge the gap between AI researchers and medical practitioners.

## 5 Discussion

The findings of this study highlight the transformative potential of artificial intelligence (AI) in the early detection and prognosis prediction of diabetic retinopathy (DR), reinforcing its role as a critical tool in modern ophthalmology. The dominance of Medicine (819 documents) and Computer Science (613 documents) in the subject area distribution underscores the interdisciplinary nature of this research, clinical needs drive technological innovation. The strong representation of Engineering (534 documents) further emphasizes the importance of developing robust diagnostic systems, particularly in medical imaging and automated screening. This convergence of disciplines suggests that successful AI applications in DR require close collaboration between clinicians, data scientists, and engineers to ensure both diagnostic accuracy and real-world usability.

The geographical distribution of research output reveals that China (487 documents) and the United States (347 documents) lead in scientific contributions, likely due to their advanced healthcare infrastructure, significant investments in AI research, and high diabetes prevalence. Meanwhile, India (279 documents) and other emerging economies demonstrate growing expertise, reflecting the global demand for cost-effective DR screening solutions. The inclusion of countries like Saudi Arabia (63 documents) and Pakistan (40 documents) indicates increasing efforts to address DR in regions with limited access to specialized ophthalmologists, aligning with the broader trend of AI-assisted telemedicine for remote screening.

Keyword co-occurrence analysis reveals that “deep learning” (link strength=12) is the most prominent AI methodology, far surpassing traditional machine learning techniques. This dominance suggests that convolutional neural networks (CNNs) and related architectures are particularly effective in analyzing retinal images for early DR signs. However, the emergence of terms like “code-free prompt” and “no-code” (strength=5) signals a shift toward user-friendly AI tools, enabling clinicians without programming expertise to leverage these technologies. This trend is crucial for widespread adoption, particularly in resource-limited settings where specialist availability is scarce.

The integration of AI into telemedicine platforms has proven especially impactful, as evidenced by studies validating smartphone-based fundus imaging combined with AI analysis for DR screening in rural and underserved areas. Such innovations address critical gaps in healthcare accessibility, reducing reliance on manual grading by specialists while maintaining high diagnostic accuracy. Additionally, economic analyses highlight the cost-effectiveness of AI-based screening, with studies demonstrating reduced healthcare expenditures through early intervention and optimized resource allocation.

Despite these advancements, challenges remain. The limited representation of alternative AI methods (e.g., support vector machines, decision trees) in the keyword analysis suggests a research gap in exploring diverse machine learning approaches beyond deep learning. Furthermore, while AI systems like IDx-DR and EyeArt have achieved regulatory approval, real-world implementation faces barriers such as data privacy concerns, algorithmic bias, and integration into existing clinical workflows. Future research should focus on explainable AI (XAI) to enhance clinician trust, as well as multi-center validation studies to ensure generalizability across diverse populations.

In conclusion, AI has revolutionized DR screening by improving diagnostic accuracy, scalability, and cost-efficiency. However, sustained progress will depend on interdisciplinary collaboration, equitable technology deployment, and continuous validation in diverse clinical settings. By addressing current limitations and fostering innovation, AI can fulfil its promise in reducing global diabetes-related blindness.

## 6 Conclusion

The integration of artificial intelligence (AI) in the early detection and prognosis prediction of diabetic retinopathy (DR) represents a transformative advancement in ophthalmology, addressing critical challenges in screening accuracy, accessibility, and cost-effectiveness. This review highlights the dominance of deep learning techniques, particularly convolutional neural networks (CNNs), in analyzing retinal images with high sensitivity and specificity, outperforming traditional manual grading methods. The interdisciplinary collaboration between medicine, computer science, and engineering underscores the synergy required to develop robust AI solutions tailored to clinical needs. Geographically, leading contributions from China, the United States, and India reflect the global push to combat DR, especially in regions with high diabetes prevalence and limited healthcare resources.

AI’s role in telemedicine has proven particularly impactful, enabling remote screening in underserved areas through portable devices and smartphone-based imaging, thereby reducing reliance on specialist availability. Economic analyses further validate the cost-saving potential of AI-driven screening, emphasizing its scalability and efficiency in high-volume settings. However, challenges such as algorithmic bias, data privacy, and the need for explainable AI (XAI) remain barriers to widespread adoption. Future research should prioritize multi-center validation studies, diverse AI methodologies beyond deep learning, and clinician-friendly tools to bridge the gap between technology and practice. By addressing these gaps, AI can fulfill its promise of reducing global diabetes-related blindness, ensuring equitable access to early intervention and improved patient outcomes.

## Data Availability

https://doi.org/10.17632/dstpngdsff.1

